# Disease-specific versus symptom-specific neuroimaging phenotypes across Alzheimer’s Disease, Parkinson’s Disease, and Small Vessel Disease

**DOI:** 10.64898/2026.11.16.26368132

**Authors:** Alberto Lazari, John Broulidakis, Grace Gillis, Gaurav Bhalerao, Daniel Halls, Fitzroy Wickham, Benjamin C. Tendler, Stamatios N. Sotiropoulos, Ludovica Griffanti, Masud Husain, Rogier B. Mars

## Abstract

**Background:** Neurodegenerative disorders are common in ageing populations, and arise from several distinct pathophysiological causes. Yet, they also share many symptoms, including cognitive and neuropsychiatric ones. It is unclear to what extent the impact of neurodegeneration on brain systems is unique or shared across disorders, and whether shared symptoms are driven by degeneration of the same brain systems.

**Methods:** We collected data from n=251 participants with Alzheimer’s Disease, Parkinson’s Disease, Small Vessel Disease, and age-matched healthy controls. All participants underwent harmonized cognitive and neuropsychiatric assessment and were scanned using the UK-Biobank MRI protocol capturing features including brain volume, white matter lesions and microstructure, tract volume, iron content, and functional connectivity.

**Results:** We replicated established case-control brain changes in Alzheimer’s Disease, Parkinson’s Disease and Small Vessel Disease compared to healthy controls. Leveraging this joint standardised dataset, we identified distinct neuroimaging features of individual neurodegenerative disorders, providing evidence for robust diagnosis-specific brain changes. Importantly, however, there was also a large degree of transdiagnostic similarity across neuroimaging markers. Testing for transdiagnostic associations between symptoms and brain phenotypes, we found that lower medial frontal pole connectivity to primary motor cortex was preferentially associated with apathy, fatigue, and depression; whereas cingulum microstructure was associated with cognitive deficits across diagnoses.

**Conclusions:** These results demonstrate that different neurodegenerative diseases are characterised by dissociable brain changes, but also common transdiagnostic and symptom-associated features. This suggests that shared symptoms across neurodegeneration may in part be driven by overlapping impact on key brain systems, such as changes in medial frontal pole connectivity and cingulum microstructure. Our work highlights that multimodal imaging biomarkers can help understand and parse symptomatic heterogeneity in neurodegenerative and ageing populations.

## Introduction

Neurodegenerative diseases are highly heterogeneous, arising from diverse pathophysiological processes (Wilson et al., 2023). Yet, they frequently share cognitive and psychiatric symptoms (Husain, 2017; Nelson et al., 2023; Bradley and Holmes, 2026). This overlap is often treated as secondary to disease-specific pathology, and most research has focused on characterising phenotypes within individual conditions. Therefore, it remains unclear whether the impact of neurodegeneration on brain systems is disease-specific or shared across diseases. It is also unknown whether the overlapping symptomatology reflects a shared disruption of specific neural mechanisms. Identifying such mechanisms underlying cognitive and psychiatric symptoms has the potential to inform the development of new therapeutic targets and enable more precise brain-based patient stratification in clinical trials.

Alzheimer’s Disease (AD), Parkinson’s Disease (PD), and Small Vessel Disease (SVD) are common conditions with high associated societal burden (Deuschl et al., 2020; Steinmetz et al., 2024; Cannistraro et al., 2019; Elmståhl et al., 2024). They have distinct pathophysiological causes and weakly correlated plasma proteomics signatures (Ali et al., 2025). With respect to brain systems, AD often impacts the hippocampi and temporal lobes (Rabinovici et al., 2008; Barnes et al., 2009; Shi et al., 2009; Frisoni et al., 2010; Jack et al., 2010; Couvy-Duchesne et al., 2025), whereas PD is primarily characterised by nigrostriatal degeneration (Kordower et al., 2013; Ulla et al., 2013; Höglinger et al., 2024; López-Aguirre et al., 2025; Madelung et al., 2025). As a prodromal phase of vascular dementia, SVD is associated with white matter abnormalities (Wardlaw et al., 2019; Duering et al., 2023). However, AD, PD and SVD patients also share several symptoms, particularly in the psychiatric domain, with symptoms such as anhedonia, apathy and depression being a common feature of all three disorders (Aarsland et al., 2009; Husain, 2017; Fang et al., 2020). Therefore, given their prevalence in the general population and their symptomatology overlap, AD, PD, and SVD are ideally suited to explore the unique and shared changes in neural systems across neurodegeneration, and whether neuropsychiatric symptoms in neurodegeneration are underpinned by overlapping effects on key brain systems. Magnetic Resonance Imaging (MRI) can be used to characterise a broad range of biological processes and brain systems *in vivo* in patients. Different MRI modalities capture complementary aspects of brain structure and function, each providing information on specific features such as brain volume, white matter microstructure, or functional connectivity (Miller et al., 2016). However, many neuroimaging studies in neurodegeneration focus on one or two imaging modalities, limiting the ability to assess the multidimensional nature of brain changes. It is increasingly being understood that integrating this multimodal information about brain changes is key for clinical translation (Markello et al., 2021; Lazari et al., 2021, 2022; Binette et al., 2025; Cherix et al., 2026).

Comparing MRI-related changes across multiple diseases often requires combining datasets from multiple sources. However, neuroimaging results are highly dependent on data acquisition and analytical choices, making it challenging to infer the presence and size of disease-specific, transdiagnostic and symptom-associated multimodal neuroimaging effects across disorders by simply pooling single-disorder and single-modality studies (e.g. through meta-analyses). Large biobanks increasingly offer a useful alternative, but they often focus on individual disorders (e.g. Alzheimer’s Disease Neuroimaging Initiative, Parkinson’s Precision Medicine Initiative) or collect large population-level data with limited phenotypic depth related to diagnosis and symptomatology (e.g. UK Biobank). As such, population studies also often cannot identify transdiagnostic and symptom-associated brain phenotypes.

To understand which neuroimaging phenotypes from the literature are unique to individual diagnoses, and which are shared or symptom-associated, a novel approach is needed, combining (a) a cohort of multiple neurodegenerative disorders and (b) multiple brain imaging modalities to facilitate an unbiased assessment of different brain imaging techniques. Here, we collected harmonized brain neuroimaging and symptom data from AD, PD, and SVD patients, and age-matched healthy controls. As part of our neuroimaging protocol, we acquired scans using an adapted UK-Biobank approach (Miller et al., 2016; Alfaro-Almagro et al., 2018; Gillis et al., 2025) which provided a range of detailed measures on brain structure, microstructure, and function.

Using these imaging phenotypes, our study had three main aims. First, we sought to establish case-control differences to healthy controls that have been previously reported in single-diagnosis, single-modality neuroimaging studies. Second, we aimed to leverage the combined multi-modal and multi-diagnostic sample to identify diagnosis-unique features capable of distinguishing individual disorders. Third, we aimed to quantify transdiagnostic overlap between disorders, and to assess symptom-associated features that may be common across diagnostic categories. To briefly anticipate our results, having established that our imaging-derived markers are able to capture diagnosis-specific changes, we found several imaging phenotypes which are shared across diagnoses and related to both psychiatric and cognitive symptoms. This demonstrates the feasibility and importance of our proposed multi-modal, multi-disorder approach to understanding neurodegenerative disease.

## Results

We collected multimodal neuroimaging data from participants with Alzheimer’s Disease (AD), Parkinson’s Disease (PD), and Small Vessel Disease (SVD), as well as from Healthy Controls (HC; Fig.1 and Fig.S1). Neuroimaging data were collected using a modified version of the UK Biobank acquisition protocol (Fig. S2), which maintained core acquisition parameters and enabled standardised preprocessing across all samples. The UK Biobank data processing pipeline (Alfaro-Almagro et al., 2018; Gillis et al., 2025) was then used to preprocess all scans and derive multimodal Imaging-Derived Phenotypes (IDPs; Fig. S3 for examples of IDPs and related distributions).

**Figure 1:**
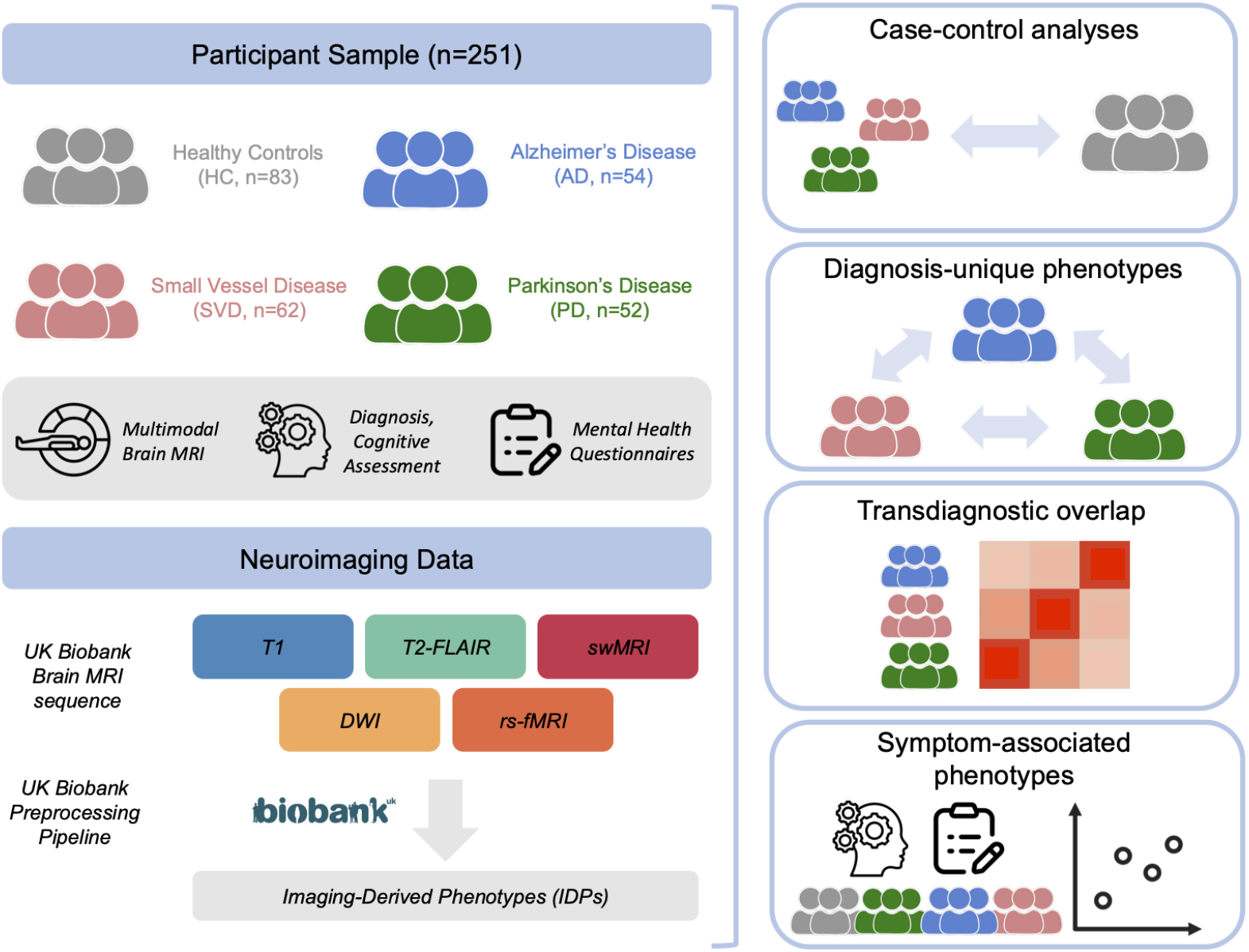
Overview of study design and analysis approach. Participant sample included Healthy Controls (HC) as well as patients with Alzheimer’s Disease (AD), Parkinson’s Disease (PD), and Small Vessel Disease (SVD). Outcome measures included multimodal neuroimaging, clinician-assigned diagnosis, cognitive assessment, and mental health questionnaires. Neuroimaging data was preprocessed using the UK Biobank Preprocessing Pipeline to generate Imaging-Derived Phenotypes. Analyses focussed on case-control comparisons; diagnosis-unique phenotypes; transdiagnostic overlap; and symptom-associated phenotypes.

### Case-control analyses

As a first step to validate our approach, we sought to establish known findings from previous case-control studies for all the neurodegenerative disorders included in this study. To control for potential confounding effects, age, sex and head size were included as covariates in all analyses. As the study spans different neurodegenerative disorders where disease severity are reflected in different symptom scores (e.g. ACE-III for AD; MDS-UPDRS for PD) and neuroimaging phenotypes (e.g. total brain volume for AD; White Matter Hyperintensities (WMHs) volume for SVD), no additional measure of disease severity was included as a confounder (Fig.2; Fig.S4).

**Figure 2:**
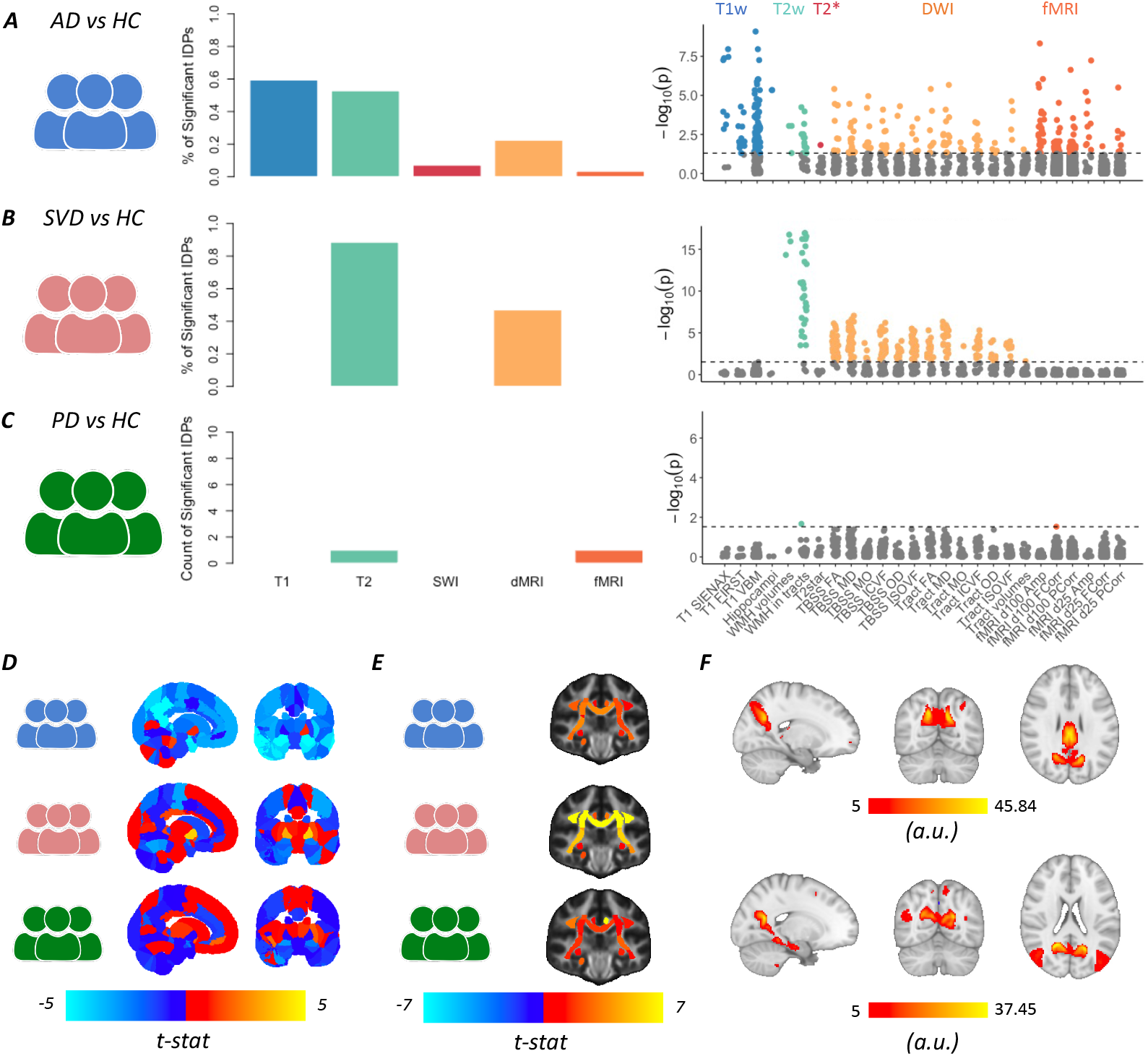
Case-controls analyses for individual neurodegenerative disorders. (A–C) Summary of significant imaging-derived phenotypes (IDPs) for each diagnosis relative to healthy controls (HC). Bar charts (left) show the proportion of significant IDPs out of the total number of IDPs within each imaging modality. Significant PD results are reported as raw counts instead of percentages. Manhattan plots (right) display significance for all IDPs ordered by modality and measure along the x-axis; IDPs surviving correction across IDPs and modalities are shown in colour, with non-significant IDPs shown in grey. (D) T-statistic maps for for T1w-derived cortical and subcortical volumes. (E) T-statistic maps for T2w-derived WMH volumes in each white matter tract. (F) Spatial maps of the two fMRI IDPs whose fluctuation amplitude has the lowest p-values in the AD-HC comparison. Abbreviations: T1 = T1-weighted imaging; swMRI = susceptibility-weighted MRI; DWI = Diffusion-Weighted Imaging; rs-fMRI = resting-state fMRI; VBM = Voxel-Based Morphometry; TBSS = Tract-Based Spatial Statistics; FA = Fractional Anisotropy MD = Mean Diffusivity; MO = Mode of Anisotropy; ICVF = Intracellular Volume Fraction; OD = Orientation Dispersion; ISOVF = Isotropic Volume Fraction; WMH = White Matter Hyperintensity; Amp = Amplitude; FCorr = Full Correlation; PCorr = Partial Correlation; d25 = 25-dimensional ICA decomposition; d100 = 100-dimensional ICA decomposition.

As a brief overview of the case-control results, we first report for each disease (AD, PD, SVD) the overall percentage of IDPs in each modality (T1w, T2w, SWI, dMRI, fMRI) that were significantly different from controls (Fig.2A-C, middle panels). A detailed breakdown of these changes (e.g. which metric in each modality is significant) is then provided through Manhattan plots (Fig.2A-C, right-most panels) and spatial maps of T1w and T2w IDPs (Fig.2D-E). The key changes found in the case-control analyses for each disease, and their relationships with previous literature, are described below.

Structurally, patients with AD (Fig.2A) had significant lowering of grey matter volume, increased WMHs, as well as reduced white matter fractional anisotropy (FA). Hippocampal volume changes, perhaps the most well-established effect in the literature (Shi et al., 2009; Gillis et al., 2025), were also evident here (tstat=−5.61 and −5.55 for right and left hippocampus respectively). Functionally, widespread changes in rs-fMRI IDPs were present: the amplitude of fluctuations in several areas was decreased, while functional connectivity had a more heterogeneous pattern, with some areas showing increased connectivity and others having decreased connectivity (Fig.S4). This is in accordance with previous literature (Badhwar et al., 2017). Although the exact location and trajectory of cortical atrophy has been debated, here we found that 60% of grey matter IDPs were significantly different from controls, pointing to a diffuse pattern of cortical atrophy. Mirroring the volumetric hippocampal changes, the two functional IDPs whose amplitude fluctuations had the greatest difference between AD patients and controls (tstat=−8.08 and −7.49 respectively) involved hippocampal and precuneus activity (Fig.2F).

SVD (Fig.2B) is characterised by white matter hyperintensities (WMHs) (Wardlaw et al., 2019). In line with this, a majority of WMH IDPs differed between SVD patients and controls. Expanding on this finding, white matter microstructure metrics, including both DTI and NODDI metrics, were significantly affected across white matter tracts; FA and MD metrics were lower and higher than controls, respectively (effect sizes for all DWI metric reported in Fig.S4). On the other hand, SVD patients did not have significant differences in volume of both grey and white matter compared to controls, although several cerebellar volume IDPs and pallidum volumes are non-significantly lower compared to controls (peak pFDR is for left pallidum, pFDR=0.03; t=3.84; cerebellar lobule VI: pFDR=0.06, t=−3.18; both not significant after Simes-based correction for testing across different modalities). Tractography-defined white matter tract volumes were not different from controls, with the exception of the forceps minor (pFDR=0.025). Taken together, these results highlight that SVD is characterised by microstructural lesions and changes in white matter, but does not present widespread macroscopic changes in grey matter volumes or white matter tracts.

Early-stage patients with PD (Fig.3C) are known to show few gross neuroimaging abnormalities. We recapitulate this finding here, as the early-stage PD patients in our cohort (UPDRS-III score mean: 25.81; SD: 9.96) had no significant volumetric differences from controls, including in subcortical areas (peak pFDR=0.23, t=−3.05 for left inferior temporal gyrus volume).The only significant IDPs for PD were WMHs in the right superior longitudinal fasciculus (pFDR=0.02, t=3.50) and one functional connectivity IDP representing connectivity between visual cortex and the dorsal attention network (pFDR=0.03, t=4.65). Some studies have highlighted iron accumulation in subcortical regions in PD, although these changes are non-linear and more evident in late-stage patients (Du et al., 2022; Ĺopez-Aguirre et al., 2025; Madelung et al., 2025). Substantia nigra and striatal regions show increasing iron accumulation with disease progression, and iron-sensitive R2* in the putamen is particularly sensitive to longitudinal changes (Ulla et al., 2013; Nikolcheva et al., 2026). Partially in line with this previous literature, we found a non-significant increase in R2* in the putamen in our early-stage PD patients compared to controls (left putamen R2*: uncorrected p=0.064, t=1.87; right putamen R2*: uncorrected p=0.070, t=1.83). In addition, white matter microstructure in internal and external capsule tracts that surround the basal ganglia was non-significantly lower compared to healthy controls (e.g. FA in the left posterior limb of the internal capsule: pFDR=0.04; t=3.56; not significant after Simes-based correction for testing across different modalities). This suggests that microstructural changes in striatal regions may be particularly sensitive to neurodegeneration in early-stage PD.

**Figure 3:**
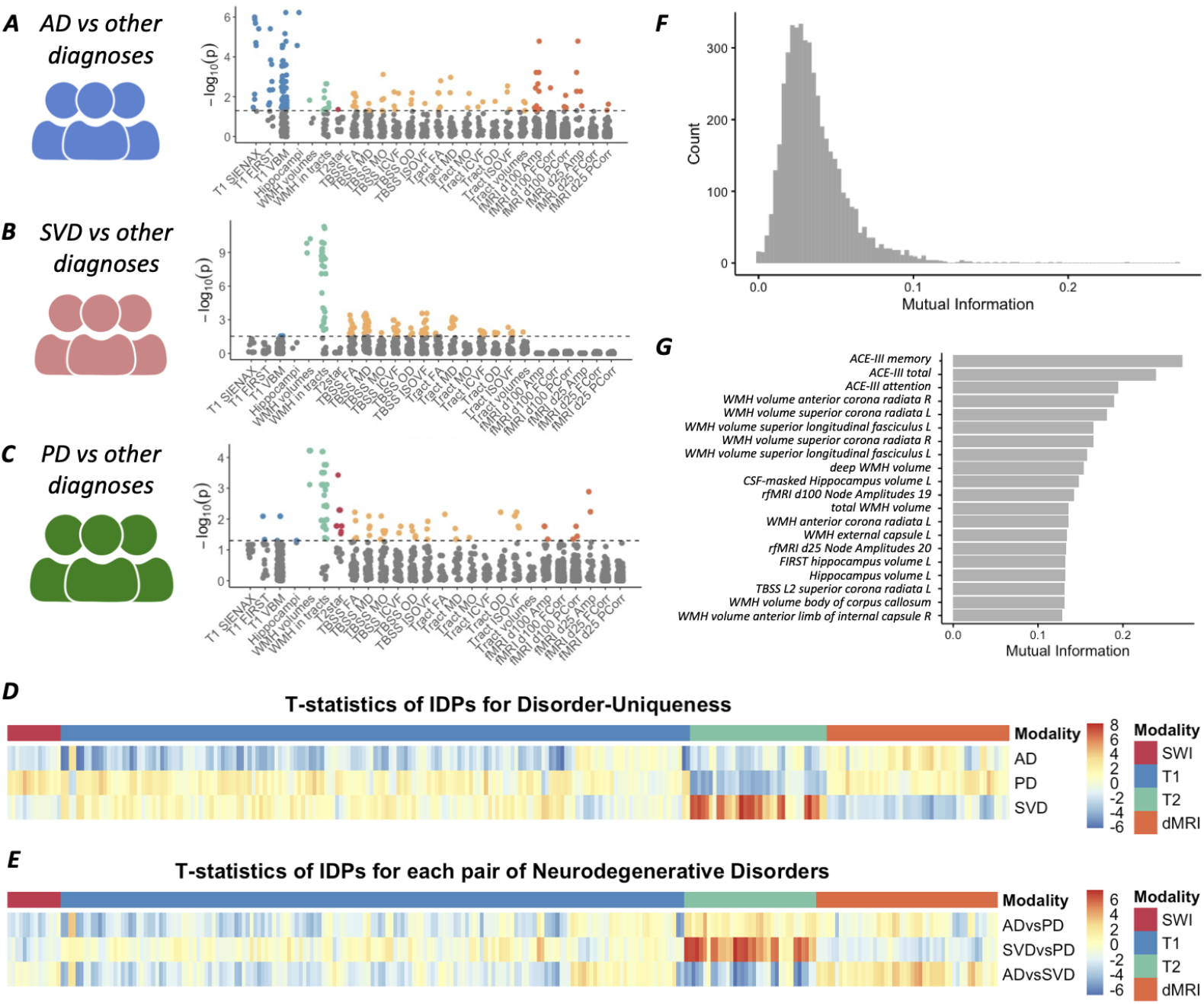
Diagnosis-unique phenotypes. A,B,C: Comparisons for all IDPs between each diagnosis and patients with other neurodegenerative diagnoses. D: Distribution of mutual information values for all IDPs and symptom measures. E. 20 measurements with the highest mutual information, and their associated mutual information value, in decreasing order. F: Heatmap of effect sizes for comparisons between each diagnosis and other neurodegenerations diagnoses. G: Heatmap of effect sizes for comparisons between each pair of diagnoses. Abbreviations: T1=T1-weighted imaging; T2=T2-FLAIR imaging; SWI=susceptibility-weighted imaging; dMRI=diffusion-weighted imaging.

As part of the preprocessing, a subset of participants (n=10) were excluded due to failed visual quality checks, outlier values on image quality metrics, or both. To test the robustness of our findings, we probe the impact that excluding these participants has on the overall results. We find that results are broadly similar when including these participants (Fig.S5).

### Diagnosis-unique neuroimaging phenotypes

Having established brain imaging features that differ from controls, we asked whether any of these features are unique to each disorder. The purpose of this analysis was to leverage our multi-diagnostic sample to identify diagnosis-unique features that are not just different from age-matched healthy controls, but also different between populations with distinct neurodegenerative conditions.

We first tested whether any IDPs differ between each diagnosis and the pooled sample of the other two diagnoses. We found that all three diseases have IDPs that are significantly different from the others (Fig. 3A-C). These results largely mirror findings from case-control analyses, with AD patients being characterised by widespread changes in multimodal imaging and SVD patients being characterised by higher volumes of WMHs and changes in diffusion-weighted metrics. Compared to AD and SVD, PD patients had fewer white matter lesions, more intact white matter microstructure, and higher grey matter volume (Fig. 3D-E).

One key difference from case-control results is that several SWI IDPs were significant for PD. In particular, R2* in the putamen was higher in PD patients compared to AD and SVD (left putamen: t=4.34, pFDR=0.00037; right putamen: t=3.42, p=0.00508). Similar to the case-control results, AD had significantly lower R2* in the left hippocampus (t=−3.00; p=0.0438). These results highlight that subcortical iron accumulation is a promising marker to distinguish patients with different neurodegenerative diagnoses.

To further probe which features may optimally distinguish the three disorders, we used a mutual information theory approach, combining neuroimaging IDPs as well as symptom data as a reference. Neuroimaging IDPs outperformed most symptom measures in distinguishing the three disorders, with the notable exception of ACE-III cognition measures (Fig. 3F-G). In addition, we found that the neuroimaging features with the highest mutual information span several brain systems and biological processes, as they included white matter lesions across many tracts, default mode network fluctuation amplitude, hippocampal volume, and corona radiata microstructure. Taken together, these analyses highlight that AD, PD and SVD show several diagnosis-specific brain phenotypes, which are identifiable across several imaging modalities.

### Transdiagnostic overlap in neuroimaging phenotypes

Having focused thus far on IDPs that distinguish patients from healthy controls and patients with different diagnostic labels from one another, we changed perspective to ask what features might be common across neurodegenerative disorders. We first tested whether the IDPs that are significant in case-control analyses at the 5% FDR threshold level are shared across AD, PD and SVD (visualized as a Venn Diagram in Fig.4A and as an UpSet plot in Fig.S6). 252 significant IDPs were unique to AD (including several resting-state and volumetric IDPs), 175 IDPs were unique to SVD (including white matter hyperintensities, white matter diffusion metrics, and volume of the forceps minor), and 2 were unique to PD (including one functional connectivity IDP representing connectivity between visual cortex and the dorsal attention network). Many IDPs were also shared across diagnoses: 8 IDPs (including WMHs in the right superior longitudinal fasciculus and diffusion metrics in internal and external capsule) were shared across AD, PD and SVD; 82 IDPs were shared across AD and SVD (including total WMH and diffusion metrics in several tracts such as the corona radiata, corpus callosum and cingulum).

**Figure 4:**
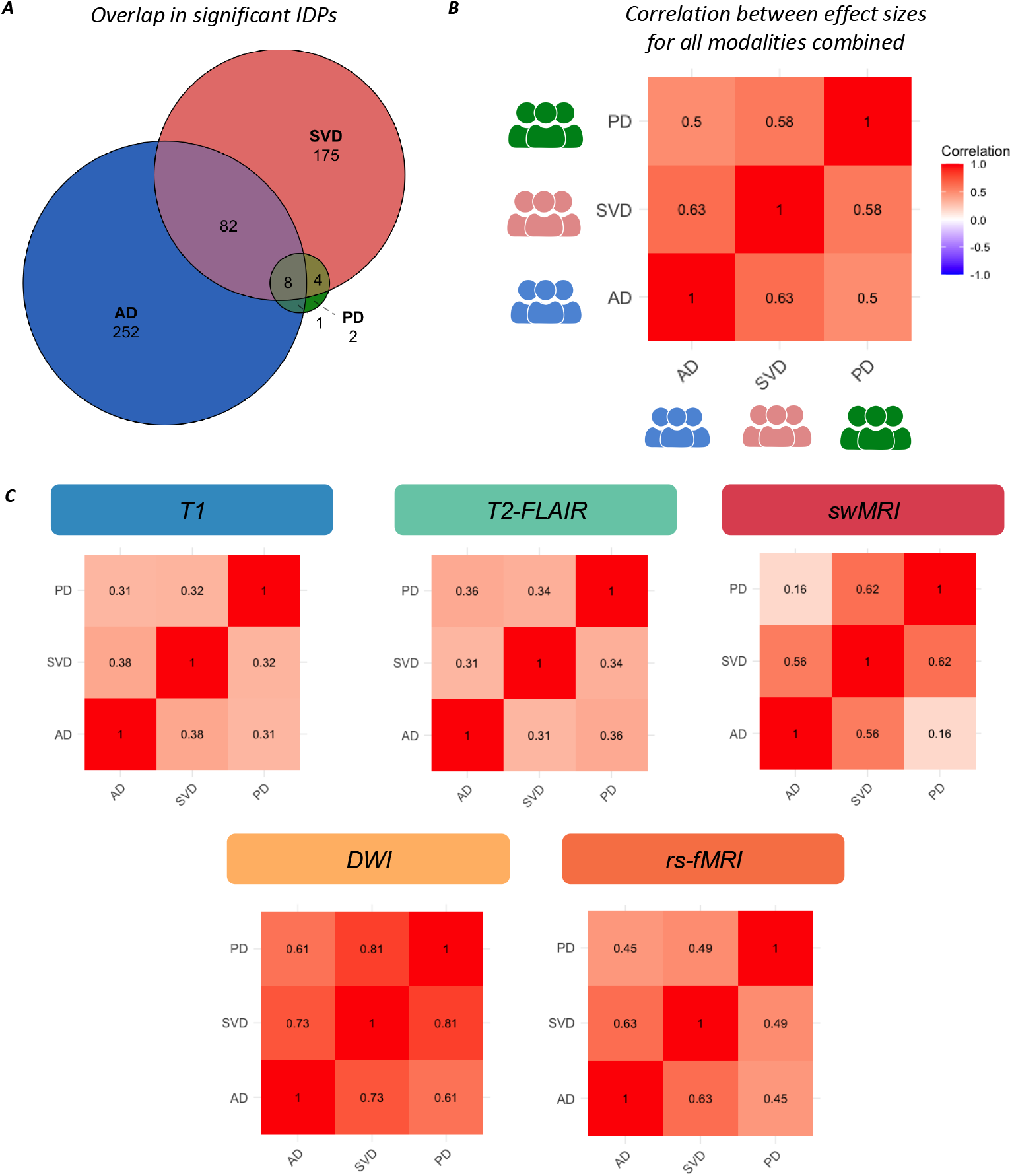
Shared phenotypes across diagnoses. A. Venn diagram of overlap between significant IDPs across diagnoses. B. Correlogram for effect size correlations when combining all IDPs across modalities. C. Correlograms for effect size correlations when analysing IDPs from each modality separately. Abbreviations: T1=T1-weighted imaging; swMRI=susceptibility-weighted MRI; DWI=Diffusion-Weighted Imaging; rs-fMRI=resting-state fMRI.

After establishing that significant IDPs overlap across diagnoses, we next examined whether their effect size distributions are also similar. Because IDPs capture measurements from different MRI modalities and spatial locations throughout the brain, comparing the distribution of effect sizes across IDPs enables us to assess whether neurodegenerative diseases exhibit a shared spatial pattern of biological alterations. To this end, we correlated effect sizes from case–control analyses across multiple MRI modalities for each pair of neurodegenerative diseases (Fig. 4B). Pooling all IDPs across modalities together, there was evidence for strong overlap in neuroanatomical changes across disorders (AD-PD: r=0.5; AD-SVD: r=0.63; PD-SVD:r=0.58). When looking at individual modalities, this overlap was strongest in DWI and fMRI IDPs (Fig.4C). For comparison, these correlations are substantially higher than similar estimates for plasma proteomics changes, which were estimated to be r=0.04 for AD-PD similarity (Ali et al., 2025). Therefore, in addition to identifying diagnosis-unique phenotypes, this analysis also uncovered a large degree of transdiagnostic overlap across neurodegenerative diagnoses. Taken together, these results demonstrate a shared spatial distribution of neuroanatomical change across diagnoses, suggesting that different pathophysiological mechanisms may converge not only on similar symptoms, but also on shared brain systems.

### Symptom-associated neuroimaging phenotypes: associations across controls and neurodegeneration patients

Given the overlap and correlation in brain changes across diagnoses, we hypothesized that some neuroimaging markers may be related to symptomatology, which is partially shared across disorders. To test this hypothesis, we combined the neuroimaging data with detailed cognitive and mental health symptomatology data. As a first step to reduce the dimensionality of the symptom data, we ran a PCA and found two key components which explain 45.46% of variance in the data (Fig.5A, Fig.5B). Principal Component 1 (PC1) loadings were high for psychiatric symptoms, including measures of depression and anxiety (BDI, GDS, HADS), anhedonia (SHAPS), apathy (AMI), fatigue (FSS, VAFS) and quality of life (WHO). Principal Component 2 (PC2) loadings were stronger for cognitive symptoms such as general cognition measures (ACE) and working memory (Digit Span). As expected, the resulting subject scores were not correlated with each other (Spearman’s test, rho=−0.012, p=0.85).

**Figure 5:**
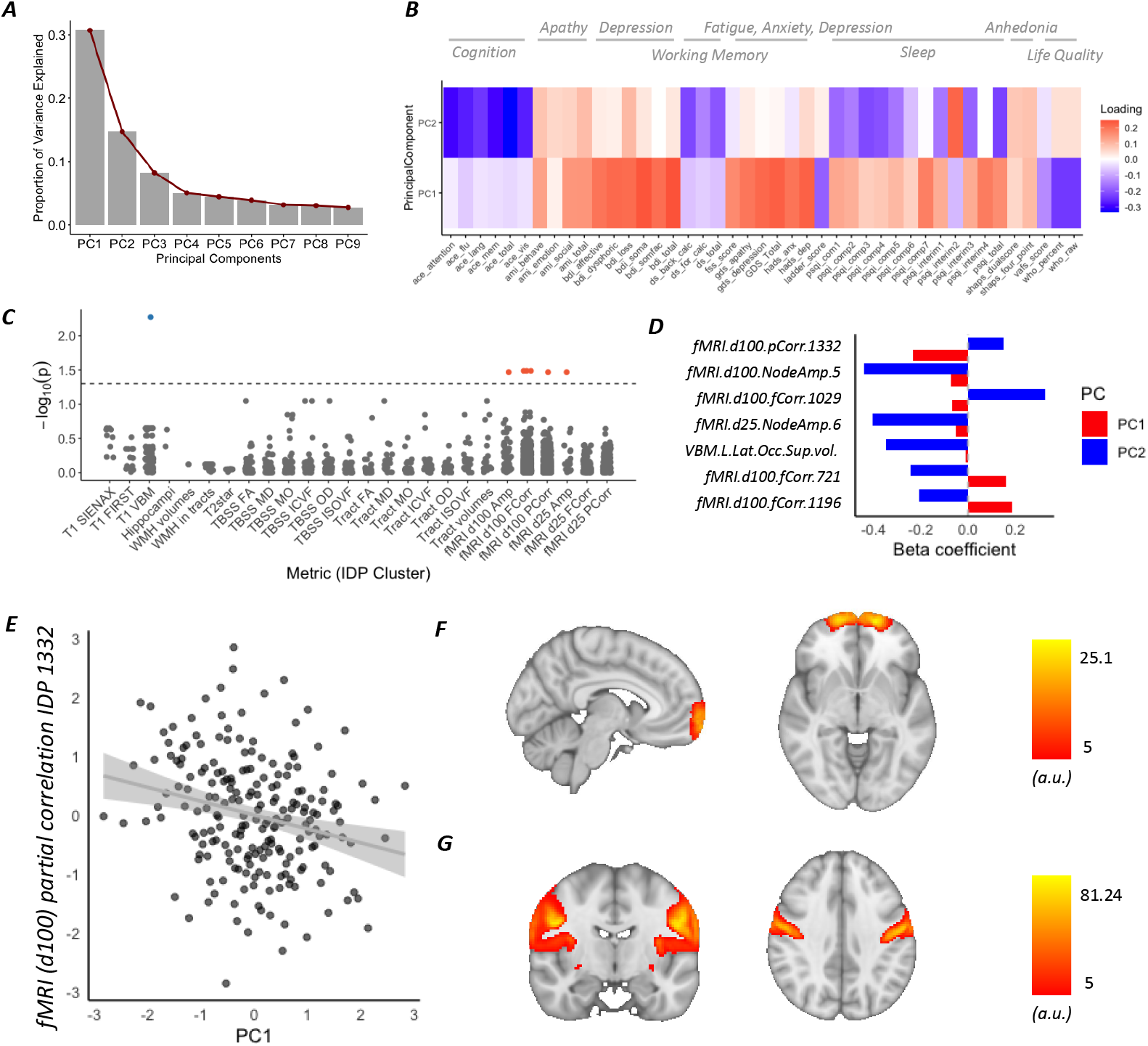
Transdiagnostic symptom-associated phenotypes across neuroimaging modalities. (A) Scree Plot for PCA of cognitive and mental health symptom data across diagnoses. (B) Heatmap of PCA loadings for PC1 and PC2 across all symptom measures, grouped by domain (Cognition, Working Memory, Apathy, Depression, Fatigue/Anxiety/Depression, Sleep, Anhedonia, Life Quality). (C,D) Beta for PC1 and PC2; results of Wald tests comparing differential associations of neuroimaging IDPs with PC1 versus PC2. (E) Association between PC1 and resting-state connectivity fMRI IDP 1332, which reflects the partial correlation between medial frontal pole and motor cortex. (F,G) Spatial maps of the fMRI networks whose partial correlation is quantified by IDP 1332. Abbreviations: BDI = Beck Depression Inventory; GDS = Geriatric Depression Scale; HADS = Hospital Anxiety and Depression Scale; SHAPS = Snaith–Hamilton Pleasure Scale, AMI = Apathy Motivation Index; FSS = Fatigue Severity Scale; VAFS = Visual Analogue Fatigue Scale; WHO-5 = World Health Organization Well-Being Index; ACE = Addenbrooke’s Cognitive Examination III; DS = Digit Span.

To explore the neural bases of transdiagnostic symptoms, we first tested whether psychiatric and cognitive symptoms are associated with any imaging marker across the entire sample of patients and healthy controls. The purpose of this analysis was to identify brain systems and biological processes involved in cognitive and mental health symptoms in neurodegenerative diseases. To this end, we pooled the entire sample to maximise the available variance in symptoms and neuroimaging data. To formally probe differences in associations with PC1 versus PC2, we applied a Wald test to all neuroimaging metrics. Several functional connectivity IDPs were differentially associated with PC1 compared to PC2 (Fig.5C, Fig.5D).

Out of the significant IDPs associated with PC1, one resting-state fMRI IDP (partial correlation between medial frontal pole and primary motor cortex) showed the strongest negative association with PC1 (*β*=−0.234; Fig.5E, Fig. 5F). This association with mental health was primarily driven by measures of apathy, fatigue, depression and anxiety (Fig.S7A). Two additional fMRI IDPs showed a significant positive association with PC1, though with lower effect sizes, both representing connectivity of the postcentral gyrus (*β*=0.161 and *β*=0.187; Fig.S8A and Fig.S8B respectively). On the other hand, several IDPs were preferentially associated with cognition as measured by PC1. In particular, lower amplitude of oscillations in the default mode network (*β*=−0.440; Fig.S8C) and volume of the lateral occipital superior gyrus (*β*=−0.349) were associated with poorer cognition; whereas higher connectivity between parietal and temporal cortex was associated with better cognition (*β*=0.327).

Taken together, this suggests that resting-state fMRI might be more sensitive than other modalities at detecting symptom-specific associations with brain phenotypes. Specifically, lower medial frontal pole connectivity to the motor network is associated with poorer mental health, but not with worse cognitive outcomes, across neurodegeneration diagnoses.

### Symptom-associated neuroimaging phenotypes: diagnosis-independent associations across neurodegenerative diseases

We then tested a related but distinct hypothesis: that any symptom-IDP association may be present independently in each individual diagnostic group. The purpose of this analysis was to identify transdiagnostic biomarkers with the potential for real-world clinical applications across heterogeneous populations with neurodegeneration. To this end, we focused on patients with neurodegenerative diagnoses, excluding healthy controls, and tested for associations that are present within each diagnostic group. No IDP was significantly associated with mental health as measured by PC1 in all diagnoses (peak pFDR=0.090 for volume of intercalcarine grey matter). In AD, PD and SVD patients, cognition as measured by PC2 was associated with microstructural integrity in the right cingulum (pFDR= 0.016; tstat=−4.29; Fig.6A-C) and resting-state functional connectivity between motor cortex and cerebellum (Fig. 6A, Fig.6D-F; pFDR=0.0058, t=5.020). When probing correlations with individual questionnaire items, lower cingulum microstructural integrity was associated with worse cognition as measured by ACE-III and Digit Span task scores (Fig.S7B), whereas higher motor cortex-cerebellum connectivity was primarily associated with better self-reported sleep measures (Fig.S7C). Taken together, this suggests that cingulum microstructure may be a promising transdiagnostic marker of cognitive decline.

**Figure 6:**
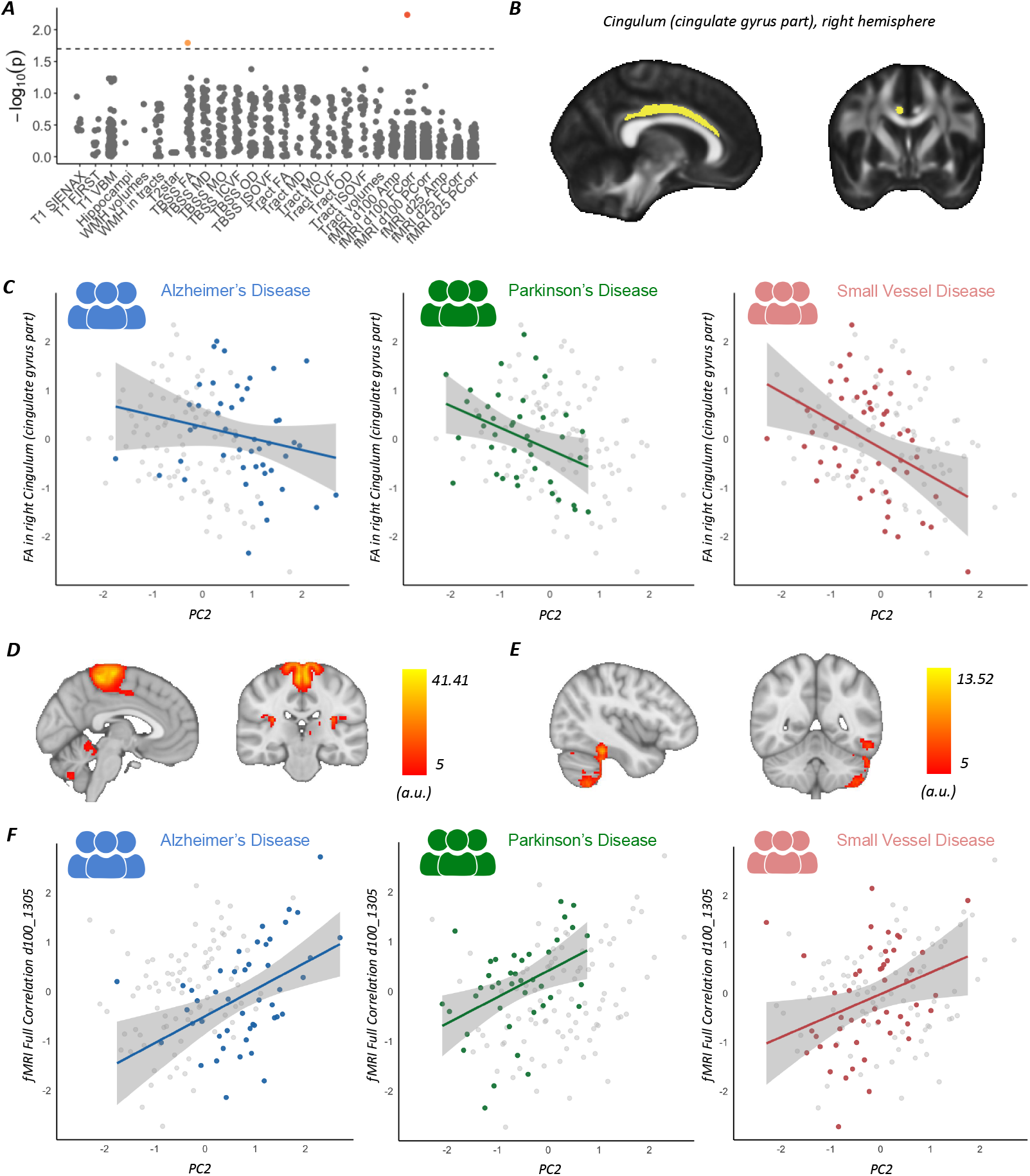
Cognitive symptoms show transdiagnostic associations with cingulum microstructure across AD, PD and SVD. (A) Manhattan plot displaying significant values for associations between PC2 scores and all imaging-derived phenotypes (IDPs), ordered by modality and measure along the x-axis. Two IDPs surviving correction are highlighted in orange: fractional anisotropy (FA) in the right cingulum (cingulate gyrus part) and an fMRI functional connectivity measure. (B) Anatomical localisation of the right cingulum (cingulate gyrus part). (C) Scatterplots showing the relationship between PC2 scores (x-axis) and FA in the right cingulum (cingulate gyrus part; y-axis) separately for AD, PD and SVD. Coloured points represent patients from diagnostic group of interest; grey points represent patients with other neurodegenerative diagnoses. (D–E) Spatial maps of the fMRI networks whose full correlation is quantified by IDP 1305. (F) Scatterplots showing the relationship between PC2 scores (x-axis) and the fMRI full correlation IDP (y-axis) separately for AD, PD and SVD. Coloured points represent patients from diagnostic group of interest; grey points represent patients with other neurodegenerative diagnoses. Abbreviations: T1 = T1-weighted imaging; swMRI = susceptibility-weighted MRI; DWI = Diffusion-Weighted Imaging; rs-fMRI = resting-state fMRI; VBM = Voxel-Based Morphometry; TBSS = Tract-Based Spatial Statistics; FA = Fractional Anisotropy MD = Mean Diffusivity; MO = Mode of Anisotropy; ICVF = Intracellular Volume Fraction; OD = Orientation Dispersion; ISOVF = Isotropic Volume Fraction; WMH = White Matter Hyperintensity; Amp = Amplitude; FCorr = Full Correlation; PCorr = Partial Correlation; d25 = 25-dimensional ICA decomposition; d100 = 100-dimensional ICA decomposition.

## Discussion

Neurodegenerative diseases in ageing populations are highly heterogeneous, but converge on common cognitive and mental health symptoms. In this study, we harnessed the shared symptomatology across three neurodegenerative conditions - AD, PD and SVD - to identify brain circuits involved in cognition and mental health in older adults. We also identified clinically relevant markers that were associated with symptomatic variability across diagnoses. By combining multimodal neuroimaging across multiple neurodegeneration diagnoses, we uncovered both disease-specific and shared brain imaging phenotypes. Overall, this work provides a comprehensive assessment of multimodal neuroimaging phenotypes in neurodegeneration.

Comparison of patients’ imaging phenotypes with healthy controls revealed key changes that were broadly in line with previous literature. Previous studies (Rabinovici et al., 2008; Barnes et al., 2009; Shi et al., 2009; Frisoni et al., 2010; Jack et al., 2010; Couvy-Duchesne et al., 2025; Kordower et al., 2013; Ulla et al., 2013; Höglinger et al., 2024; López-Aguirre et al., 2025; Madelung et al., 2025; Wardlaw et al., 2019) have suggested that AD, PD and SVD are associated with distinct diagnosis-specific changes in several imaging modalities, but few studies have directly compared these different patient populations using standardised neuroimaging approaches. Here, we provided evidence that individual neurodegenerative disorders exhibit distinct phenotypes, observable through multimodal neuroimaging.

Beyond diagnosis-specific phenotypes, we observed substantial overlap between disorders. A plausible explanation is that while the origins of pathology differ, these processes converge on common functional pathways (Wilson et al., 2023). This may account for the greater AD-PD overlap detected in our neuroimaging study compared with previous proteomic analyses, which reported limited overlap (Ali et al., 2025). Such an explanation is also compatible with the finding that white matter microstructure IDPs exhibited the greatest overlap across disorders, since they are sensitive to a broad range of biological changes affecting myelinated long-range projections (Lazari et al., 2018; Mancini et al., 2020; Lazari and Lipp, 2021). Rather than reflecting only diagnosis-specific pathology, alterations in white matter microstructure may therefore represent a common marker of disease progression (Peter et al., 2025; Binette et al., 2025). It has long been hypothesised that common cognitive and psychiatric symptoms in different neurodegenerative diseases might arise from shared disruption of common neural mechanisms (Husain, 2017; Cayir et al., 2026; Bradley and Holmes, 2026). Here, we found evidence in support of this proposal. In particular, medial frontal pole connectivity with primary motor cortex was lower in participants with worse mental health symptoms (Fig.5), while lower cingulum microstructure integrity was associated with poorer cognition across neurodegenerative disorders (Fig.6).

Medial frontal pole cortex is well established as a key site for reward processing and value-based decision making (Passingham et al., 2010; Neubert et al., 2015), and has historically been linked to psychopathology (Bludau et al., 2016; Fettes et al., 2018). More recently, primary sensory and motor cortices have also been implicated in depression and anxiety (Zhang et al., 2024; Hamilton et al., 2026), including from a previous study examining transdiagnostic neural correlates of psychopathology (Kebets et al., 2019). In this context, our study provides a link between these two literatures, by suggesting that in ageing populations, the connectivity between medial frontal pole and primary motor cortex is involved in several aspects of psychopathology, including apathy, depression and anxiety.

We found that lower integrity of cingulum microstructure was associated with poorer cognition across diagnoses. This correlation was driven by ACE-III and digit span scores, indicating a robust association with cognition. It was also present across AD, PD and SVD, suggesting that cingulum FA can be used as a scalable biomarker in clinical trials aiming to characterise and alter cognitive trajectories in neurodegeneration. This result matches previous findings in individual AD and SVD cohorts (Bozzali et al., 2012; van der Holst et al., 2013; Hirschfeld et al., 2023), including using a similar UK Biobank neuroimaging protocol (Gillis et al., 2025), and extends them to a transdiagnostic sample spanning multiple neurodegeneration diagnoses.

Biomarkers play a growing role in patient enrichment and stratification for clinical trials in neurodegeneration. Blood-based and PET-based markers (Hampel et al., 2023; Jack Jr et al., 2024) are increasingly being used to select patients more accurately by pathophysiology and disease stage. However, MRI-based neuroimaging also has the potential to support future neurodegeneration trials, in particular to complement other biomarker modalities, as each modality provides only a partial picture (Jack et al., 2013; Vemuri et al., 2015; Grande et al., 2025). For example, it has recently been shown that, for the same PET-based biomarker profile, more clinically advanced AD patients show greater WMH burden (Binette et al., 2025). Our results show that fMRI can reveal symptom-specific associations, in agreement with previous connectivity-behaviour associations (Kebets et al., 2019; Elliott et al., 2018; Park et al., 2020), whereas structural MRI biomarkers (e.g. T1w, T2w, and diffusion MRI features) show stronger differences between diagnoses. The results further reinforce the idea that imaging markers can be valuable in clinical trials of neurodegeneration, but need to be chosen carefully depending on trial design and objectives.

This study also represents a methodological advancement, as it provides one of the first applications of a UK Biobank-style protocol - i.e. a relatively brief, scalable, high-resolution and multimodal acquisition protocol - to study a clinical neurodegeneration cohort. Building on a previous study which demonstrated the feasibility of the protocol in the context of a memory clinic (Gillis et al., 2025), the current investigation shows that UK Biobank-style multimodal MRI protocols can be adapted for use in clinical research across neurodegenerative disorders. This suggests that it may be possible to harmonise neuroimaging protocols between smaller cohorts and larger biobanks, potentially facilitating their future integration (He et al., 2022).

Revealing which neuroimaging markers are unique, shared or symptom-associated across neurodegenerative disorders also holds important implications for reverse translation to preclinical studies. Rodent models of neurodegenerative diseases are fundamental for probing the efficacy potential of novel therapies. Recent work has opened the door to quantitatively assessing cross-species similarities in neuroanatomy between rodents and humans (Beauchamp et al., 2022; Mandino et al., 2022; Pagani et al., 2023; Gutierrez-Barragan et al., 2024; Lazari et al., 2024; Jaroszynski et al., 2025). However, mouse models often do not have a 1-to-1 relationship with human diseases (Dawson et al., 2018), and we still lack an understanding of which mouse model may be best for recapitulating different features of multi-faceted neurodegenerative disorders. In this respect, multimodal neuroimaging offers a promising approach to systematically capturing the phenotypic complexity of neurodegeneration in both humans and mouse models, enabling a more quantitative link between patient cohorts and preclinical disease models.

Finally, this study used a cross-sectional design, which carries inherent limitations. Given the time-varying nature of both symptomatology and neurodegeneration patterns in many disorders (Vogel et al., 2021; Joza et al., 2025), symptom-neuroimaging associations are likely to differ when investigated at earlier pre-symptomatic stages, or later more advanced ones. Therefore, longitudinal studies may be needed to supplement these results and validate whether similar associations are present across different stages of neurodegeneration. Moreover, our approach focussed on patients who recently sought clinical care. This limits its applicability to broader subclinical patient populations, but is more likely to provide translatability to other clinical research settings such as clinical trials. A further caveat is that fMRI signals are modulated by both neuronal activity and neurovascular coupling; therefore, it is possible that our fMRI results may be driven by vascular changes associated with ageing and neurodegeneration (Nortley et al., 2019; Shaw et al., 2021; Bonnar et al., 2023). In sum, our findings on symptom-associated phenotypes across neurodegenerative diagnoses open the door to further transdiagnostic studies in neurodegeneration. While transdiagnostic approaches have become commonplace in psychiatric research (Gillan et al., 2016; Dalgleish et al., 2020; Nord et al., 2021; Nord and Garfinkel, 2022; Townend et al., 2025), yielding many advances in our understanding of mental health conditions, they are still underused in neurology. Given the large amount of symptomatic overlap between patients with different neurodegenerative diseases, this may be a promising avenue for further research into the neurobiological bases of a wide range of symptoms such as apathy and anhedonia (Husain, 2017; Husain and Roiser, 2018).

## Methods

### Participants

251 participants were recruited as part of this study (n=54 AD; n=52 PD; n=62 SVD; n=83 HC). Patients were referred to neurology examination due to cognitive complaints and were diagnosed following standard care pathways in Oxfordshire. Demographic data including age and sex for each group is described in Supplementary Table S1.

Exclusion criteria included the presence of multiple neurodegenerative diagnoses, or the diagnosis of additional neurological disorders such as stroke. Since structural scans are needed for further preprocessing and registration of other neuroimaging modalities, participants lacking full T1-weighted or T2-weighted scans were not included in the final sample for the study. In addition, three participants lacked Diffusion-Weighted Imaging and one participant lacked functional MRI scans. Thirty-three participants lacked full symptomatology data, and were thus excluded from symptom-association analyses. Sample size is comparable to previous studies in neurodegeneration and in line with conservative sample size estimates for cross-sectional studies linking behavioural metrics to multimodal neuroimaging metrics (Lazari et al., 2021).

Before starting the study, all participants gave written informed consent. Ethics approval was granted by the relevant local ethics committee (IRAS ID: 341156. Ethics Approval Reference: 24/EE/0173). All aspects of the study were carried out according to the Declaration of Helsinki.

### Clinical and Questionnaire Data

All patients underwent the same evaluation for cognitive and mental health symptoms. This included the Addenbrooke’s Cognitive Examination–III (ACE-III) and the Digit Span task (DS; forward and backward). It also included the following questionnaires: Apathy Motivation Index (AMI); Snaith-Hamilton Pleasure Scale(SHAPS); Geriatric Depression Scale (GDS); Beck Depression Inventory (BDI); Hospital Anxiety and Depression Scale (HADS); Fatigue Severity Scale (FSS); Visual Analogue Fatigue Scale (VAFS); Pittsburgh Sleep Quality Index (PSQI); WHO Quality of Life assessment; and the Cantril Ladder Scale. Clinical and Questionnaire evaluations were collected together as part of a single data collection session and separately from MRI data collection (median interval between clinical/questionnaire and MRI data collection was 43 days).

### Acquisition of Multimodal MRI Data

The full MRI acquisition protocol is available online: https://dx.doi.org/10.5287/ora-xvkz4kvym. The MRI protocol was set up to closely follow acquisition parameters from the UK Biobank brain imaging protocol (Miller et al., 2016) on a 3T Siemens Magnetom Verio scanner at the John Radcliffe Hospital (Oxford, United Kingdom). Key adaptations included the removal of task fMRI sequences to minimise scanning time and ensure feasibility across the diverse patient populations involved in the study. The protocol modalities and parameters were as follows:

– T1-weighted imaging: an MPRAGE sequence with TR = 2000 ms, TE = 2.01 ms, TI = 880 ms. Resolution was 1 mm isotropic. Flip angle was 8 degrees.
– T2-weighted imaging: a fluid-attenuated inversion recovery (T2-FLAIR) sequence with TR = 5000 ms, TE = 397 ms, TI = 1800 ms. Resolution was 1.05 mm × 1 × 1 mm.
– Susceptibility-weighted imaging (SWI): a scan with resolution of 0.9× 0.9 × 3 mm; matrix size of 48 x 22 4x 256; TE1/TE2 = 9.42/17.89 ms; TR = 27 ms. Data was saved after combining across receive coils.
– Diffusion-weighted imaging (DWI): echo planar imaging (EPI) was acquired with multiband factor = 3, TR = 3600 ms, TE = 92 ms, and resolution = 2.0 mm isotropic. Anterior–posterior (AP) and posterior–anterior (PA) phase-encoding directions were used. The AP scan sequence consisted of 108 diffusion directions: 8 volumes with b = 0 s/mm^2^, 50 with b = 1000 s/mm^2^, and 50 with b = 2000 s/mm^2^. The PA scan consisted of 6 directions (4 b = 0 s/mm^2^ and 2 with b = 1000 s/mm^2^), to support distortion correction.
– Resting-state functional MRI (rs-fMRI): echo planar imaging (EPI) was acquired with multiband factor = 8, TR = 735 ms, TE = 39 ms, flip angle = 52 degrees, resolution = 2.4 mm isotropic.

### Preprocessing of Multimodal MRI Data

The MRI data was preprocessed using a modified version of the UK Biobank image analysis pipeline (Alfaro-Almagro et al., 2018), with additional modifications as previously reported (Griffanti et al., 2022; Gillis et al., 2025).

In brief, the data was denoised using well-established FSL-based (Smith et al., 2004; Jenkinson et al., 2012) modality-specific tools and was then registered to MNI space using procedures that have been developed to be robust across the entire UK Biobank sample. For T1-weighted scans, the pipeline applied gradient distortion correction, defacing, brain extraction, affine and non-linear registration to standard space. T1-weighted scans were then used as initial reference for registration across all modalities. Gradient distortion correction (GDC) was applied early in all modalities to remove spatial warping from gradient nonlinearities; a fieldmap was derived from additional b=0 images within the DWI acquisition.

After preprocessing and registration, Imaging-Derived Phenotypes (IDPs) were extracted. IDPs are subject-wise metrics of key features from each imaging modalities. A standard list of UK Biobank IDPs was extract for each modality. Additional dementia-relevant IDPs were also estimated for each subject, as previously described (Griffanti et al., 2022; Gillis et al., 2025). In particular, we focussed on extracting white matter tract volumes, tract-wise WMH volumes, and lesion-masked CSF-masked volumetric IDPs.

### MRI Quality Control

Quality control was carried out using both visual and data-driven checks. First, raw scans and template-registered scans were visually inspected. Second, as part of the preprocessing pipeline, several automatic QC metrics were extracted, such as contrast-to-noise ratio and discrepancy between each modality and template values (full metrics list described here: (Gillis et al., 2025)). To ensure QC-based exclusions were not biased towards a specific diagnostic group, we used distribution-based cut-offs (using the interquartile range of automated QC metrics) rather than absolute thresholds to determine participant exclusions.

Two participants were found to have gross neuroanatomical abnormalities; three others were excluded as part of the other quality checks. Two, three and six participants were excluded because they were outliers on measures of T1-weighted contrast-to-noise ratio, signal-to-noise ratio and template discrepancy, respectively. The overlap between these exclusions meant a total of 10 participants were excluded from the analyses, in line with previous studies (Gillis et al., 2025). Given the low number of participants failing quality checks, we validated key analyses both including and excluding the low-quality participants in the dataset.

### Main Statistical Analyses

Continuous variables were normalised to rank-based inverse normal transformation, as implemented in the bestNormalize R package (Peterson, 2021). Such variables included all IDPs, symptoms scores, age and head size. For each analysis, a linear model was fit to each IDP in order to estimate p-values, t-statistics and beta coefficients. In line with previous adaptations of UK Biobank analyses to clinical samples, we only used key covariates of age, sex and head size in our models (Gillis et al., 2025).

For the case-control analyses, the following linear model was fit to the data, including only healthy controls and one diagnostic category at a time:

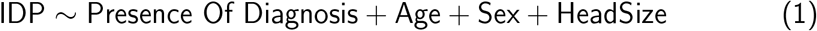

For the diagnosis-unique analyses, the following linear model was used, including relevant diagnostic categories but no healthy controls:

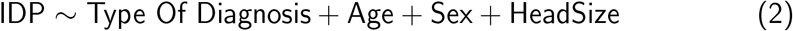

For the symptom-association analyses, the following linear model was fit to the entire sample, taking care to correct for Diagnosis when principal component subject scores different between diagnostic categories:

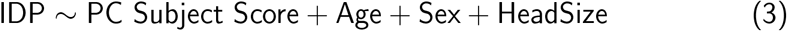

A False Discovery Rate (FDR) approach was used to correct for multiple comparisons within each modality, with within-modality corrected p-values referred to as pFDR (Yekutieli, 2008). For more rigorous correction for multiple comparisons across modalities, a two-step approach using FDR was used, following previous literature (Gillis et al., 2025). In the first step, a Simes test was used to calculate modality-wise p-values. In the second step, IDPs from significant modalities at the 5% FDR threshold level were tested with FDR at a more stringent threshold, which was adjusted according to the number of significant modalities from the first step. This procedure ensured that p-values are corrected not only for multiple comparisons within the entire modality (e.g. correcting for all T1w IDPs), but also for multiple comparisons across modalities (e.g. correcting for all T1w, T2w, SWI, DWI and fMRI IDPs tested in a given model). For analyses of overlap in significant IDPs across diagnoses, significance was based on FDR-corrected p-values without application of the Simes test; using a common significance criterion across diagnoses facilitated direct comparisons of overlapping IDPs. T-statistics were reported as the primary measure of effect size. For analyses using the two-step approach to Wald tests, beta coefficients were reported instead, as they directly reflect the underlying parameter estimates used in the test.

### Diagnosis-unique phenotypes

To identify diagnosis-specific phenotypes, linear models are applied as described above, contrasting each diagnostic group against all other diagnoses individually, as well as against the combined set of all remaining groups. To complement this targeted analysis with a unified whole-dataset assessment of how diagnosis-unique individual IDPs and clinical variables are, we conduct a mutual information (MI) analysis using the R package *infotheo*. Continuous variables are discretized into five equiprobable bins using equal-frequency binning. MI is then computed between each predictor and diagnostic label, with values estimated independently for all available predictors.

### Additional Statistical Analyses

Principal Component Analysis (PCA) and Linear Models were run using the *prcomp* and *lm* functions in the *stats* package in R. The PCA output was then used to extract participant scores for the first two principal components. Normalization of IDPs was run using the *step orderNorm* function in the *recipes* library in R.

## Data Availability

The full MRI acquisition protocol is available online: \url{https://dx.doi.org/10.5287/ora-xvkz4kvym}. Code to produce all results described in the manuscript will be openly shared upon publication.

## Acknowledgements

This work was supported by a BBSRC grant [BB/X013227/1] and an MRC Fellowship [MR/Y010698/1] granted to RBM; by an ERC Consolidator award [ERC 101000969] granted to SNS; and by the NIHR Oxford Health Biomedical Research Centre (NIHR203316). The views expressed are those of the author(s) and not necessarily those of the NIHR or the Department of Health and Social Care. The Centre for Integrative Neuroimaging was supported by core funding from the Wellcome Trust (203139/Z/16/Z). For the purpose of Open Access, the author has applied a CC BY public copyright license to any Author Accepted Manuscript version arising from this submission.

## Data Sharing

The full MRI acquisition protocol is available online: https://dx.doi.org/10.5287/ora-xvkz4kvym. Code to produce all results described in the manuscript will be openly shared upon publication.

## Supplementary Information

**Figure S1.**
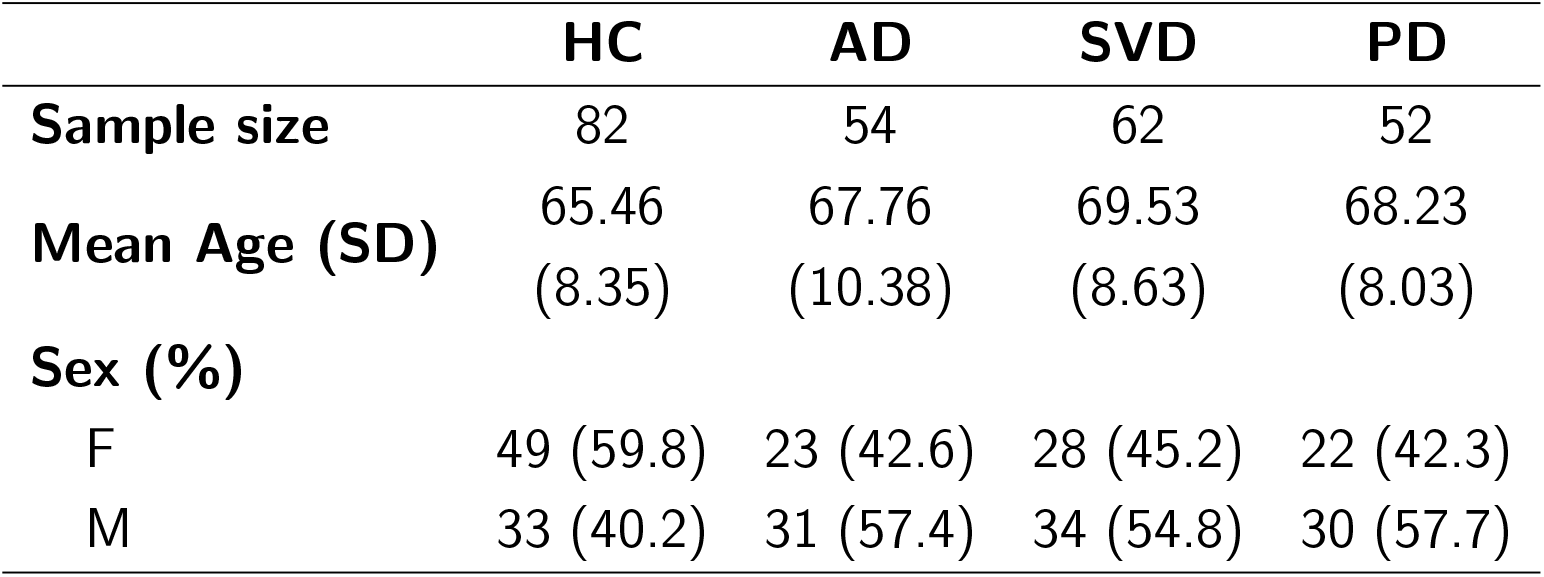
Demographics Table. Characteristics of patient sample for age-matched Healthy Controls (HC), Alzheimer’s Disease (AD), Parkinson’s Disease (PD), and Small Vessel Disease (SVD).

**Figure S2.**
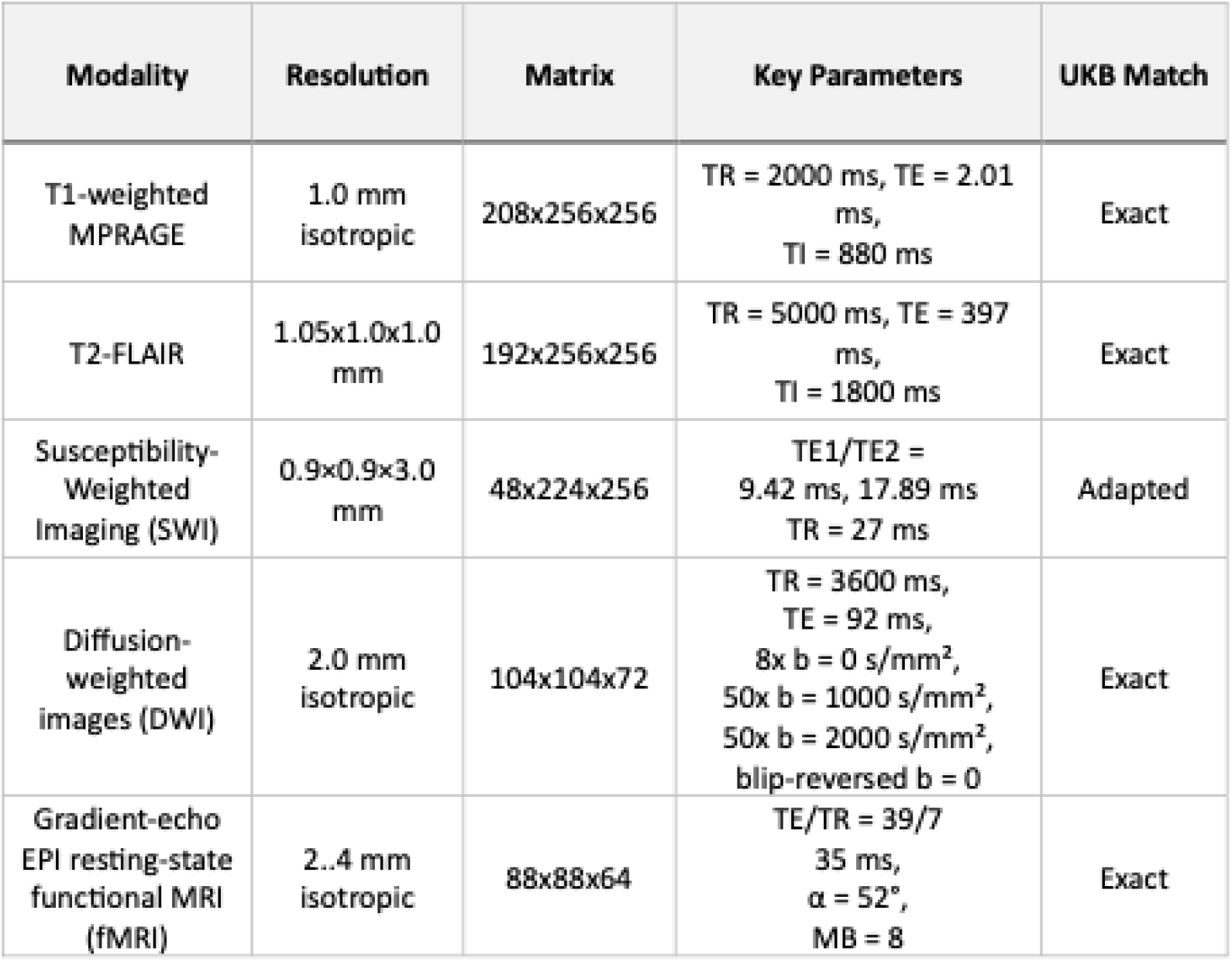
MRI Protocol summary table and comparison to UK Biobank. The MRI sequence used for the UK Biobank was applied in this study for use in clinical research. The majority of sequences were set up to exactly match the original protocol. Susceptibility-weighted imaging (SWI) was adapted to match the resolution of the UK Biobank SWI protocol using sequences available.

**Fig. S3.**
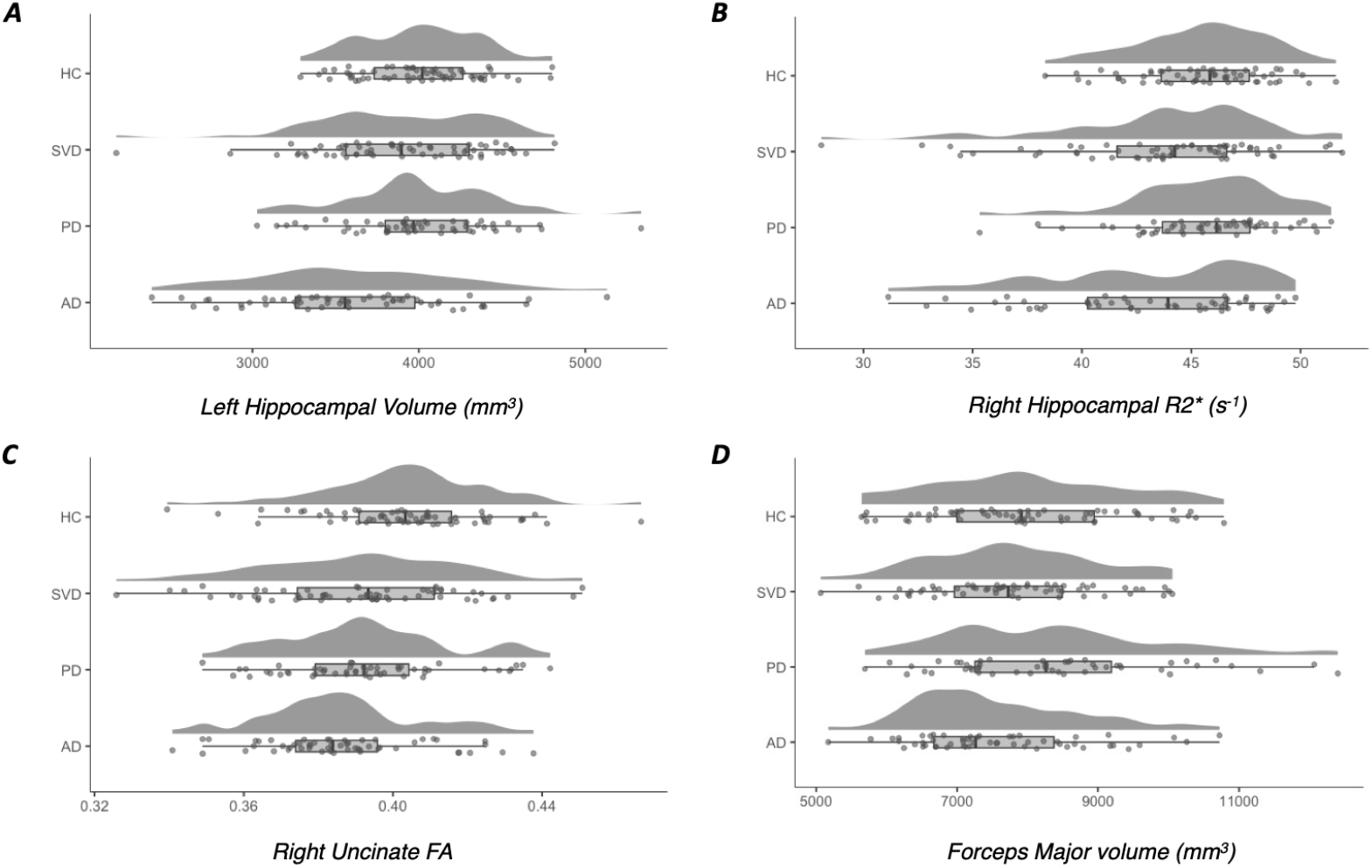
Raincloud plot by diagnosis of four IDPs for illustrative purposes. (A) Left Hippocampal Volume, (B) Right Hippocampal R2*, (C) Right Uncinate Fasciculus FA, and (D) Forceps Major volume.

**Fig. S4.**
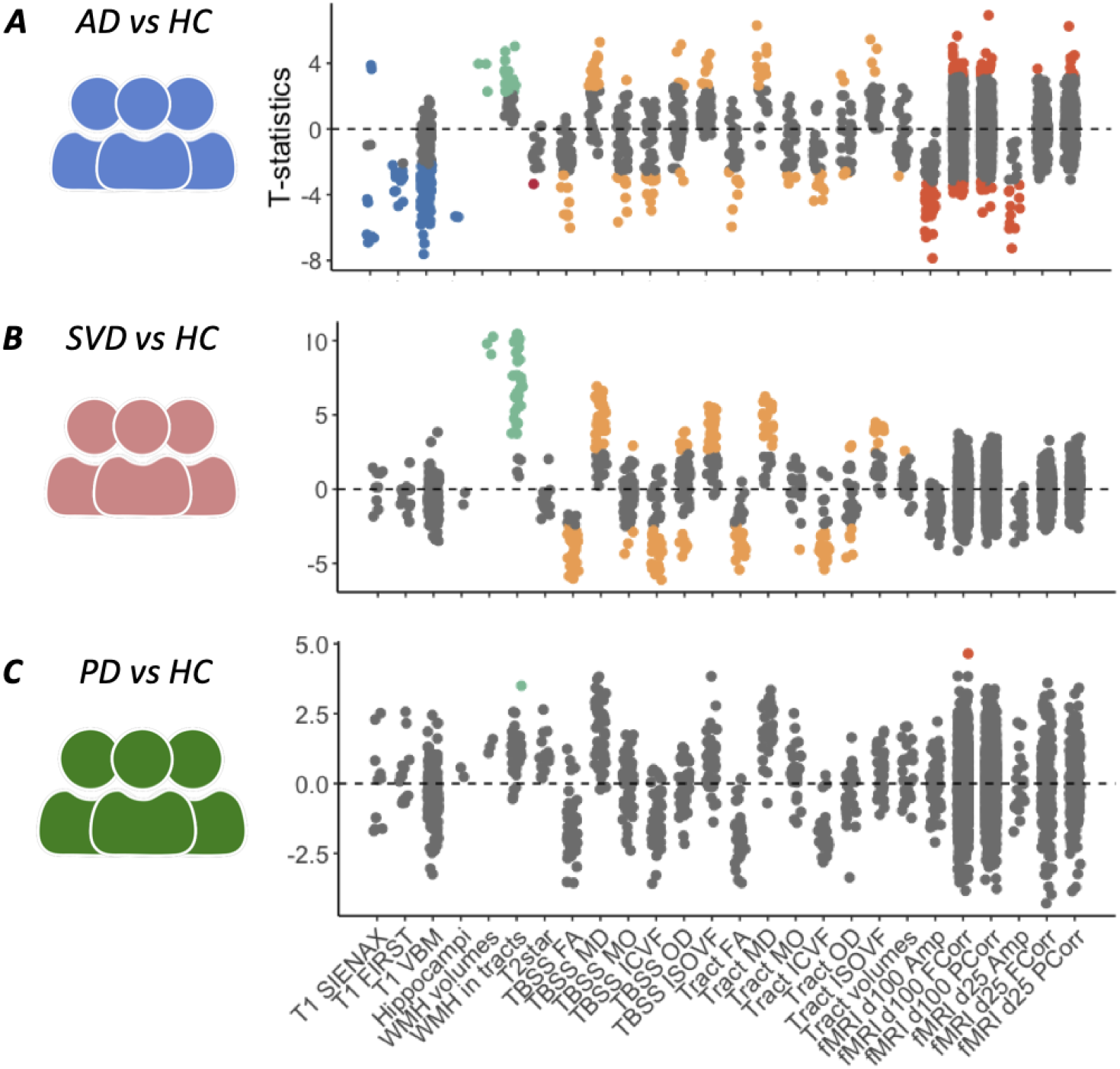
Manhattan Plot of effect sizes for case-control comparisons. Abbreviations: T1 = T1-weighted imaging; swMRI = susceptibility-weighted MRI; DWI = Diffusion-Weighted Imaging; rs-fMRI = resting-state fMRI; VBM = Voxel-Based Morphometry; TBSS = Tract-Based Spatial Statistics; FA = Fractional Anisotropy MD = Mean Diffusivity; MO = Mode of Anisotropy; ICVF = Intracellular Volume Fraction; OD = Orientation Dispersion; ISOVF = Isotropic Volume Fraction; WMH = White Matter Hyperintensity; Amp = Amplitude; FCorr = Full Correlation; PCorr = Partial Correlation; d25 = 25-dimensional ICA decomposition; d100 = 100-dimensional ICA decomposition.

**Fig. S5.**
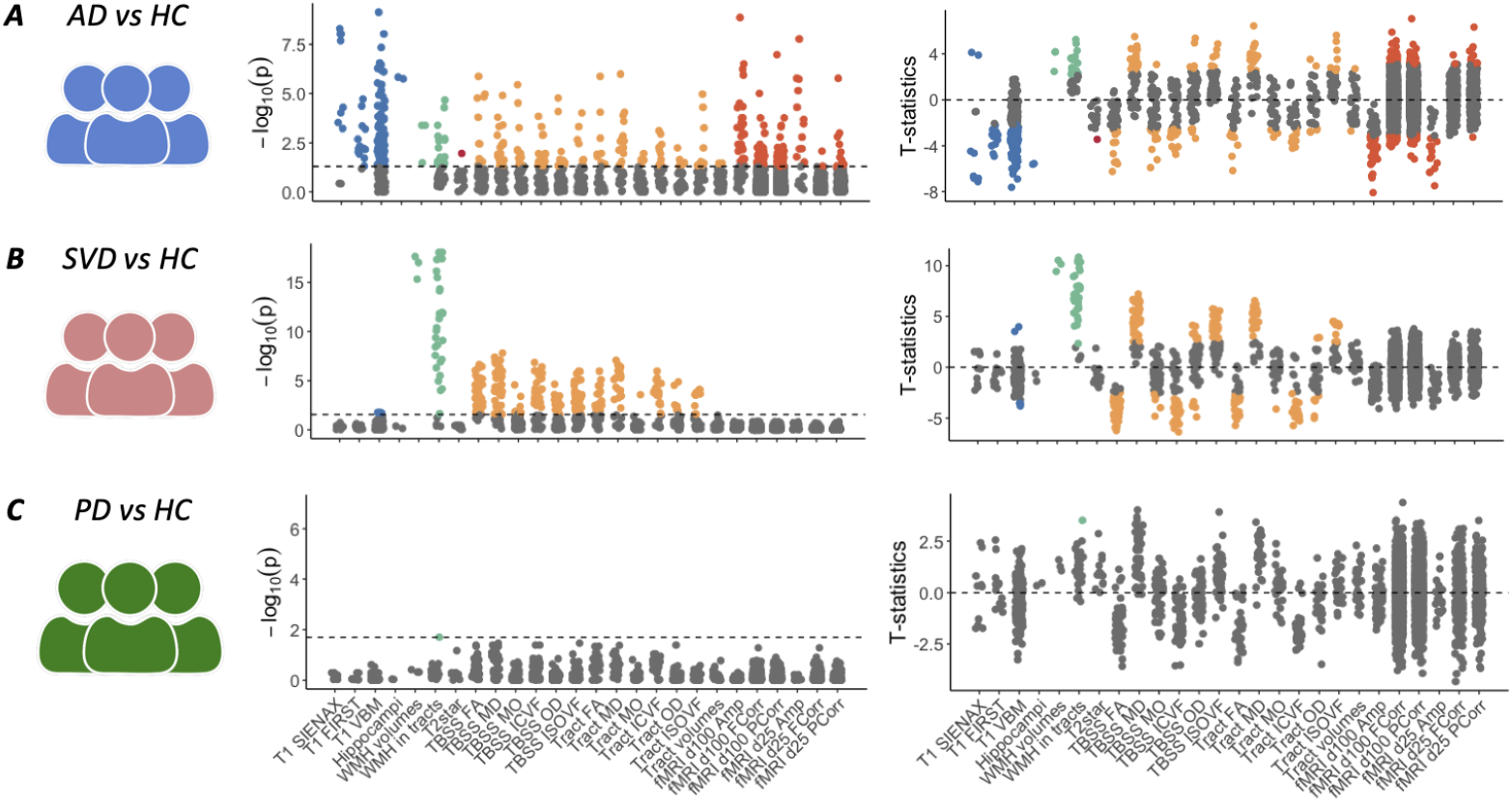
Case-control differences in multimodal MRI markers after inclusion of participants discarded during QC. Abbreviations: T1 = T1-weighted imaging; swMRI = susceptibility-weighted MRI; DWI = Diffusion-Weighted Imaging; rs-fMRI = resting-state fMRI; VBM = Voxel-Based Morphometry; TBSS = Tract-Based Spatial Statistics; FA = Fractional Anisotropy MD = Mean Diffusivity; MO = Mode of Anisotropy; ICVF = Intracellular Volume Fraction; OD = Orientation Dispersion; ISOVF = Isotropic Volume Fraction; WMH = White Matter Hyperintensity; Amp = Amplitude; FCorr = Full Correlation; PCorr = Partial Correlation; d25 = 25-dimensional ICA decomposition; d100 = 100-dimensional ICA decomposition.

**Fig. S6.**
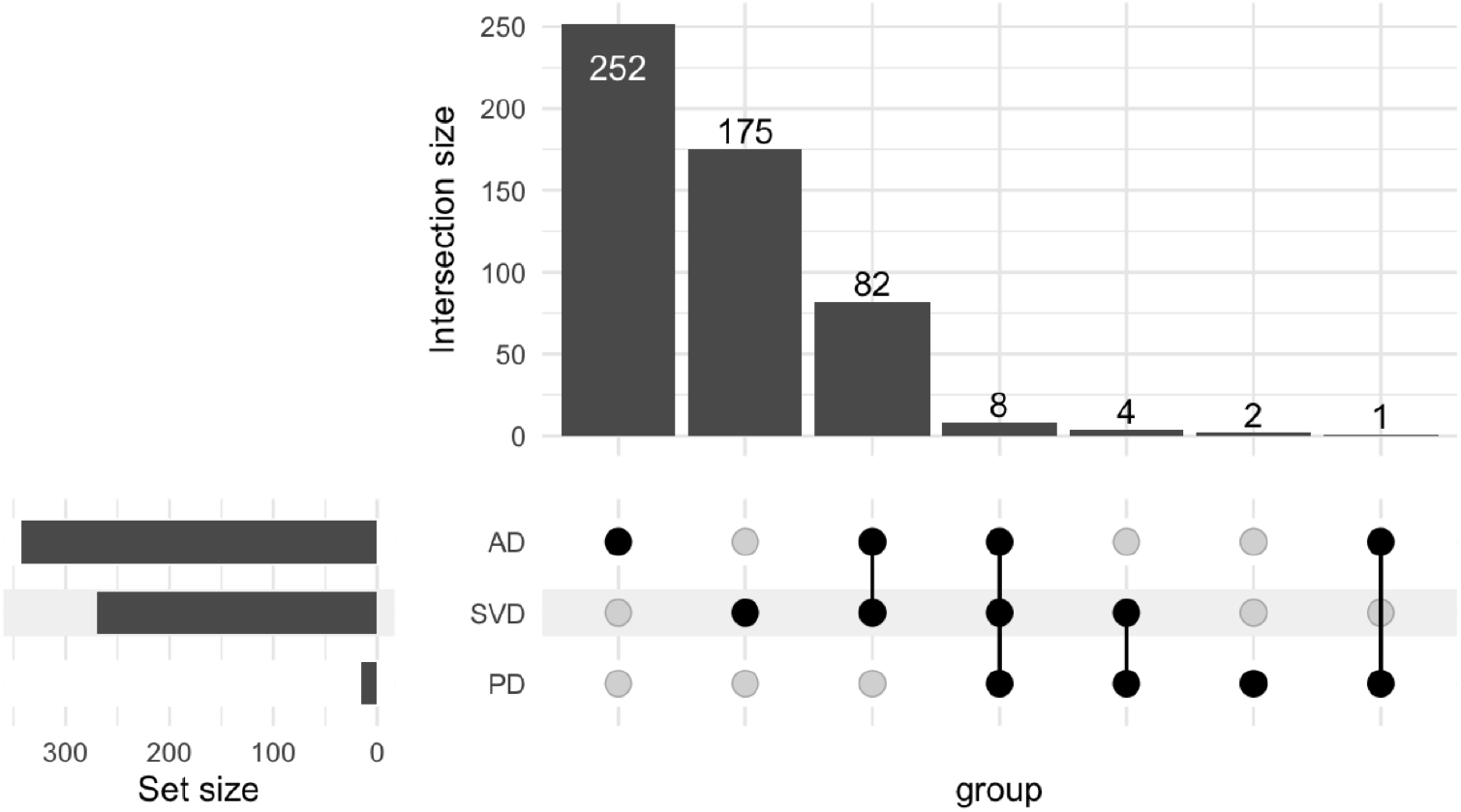
UpSet plot of significant IDPs for AD, PD and SVD and their intersections.

**Fig. S7.**
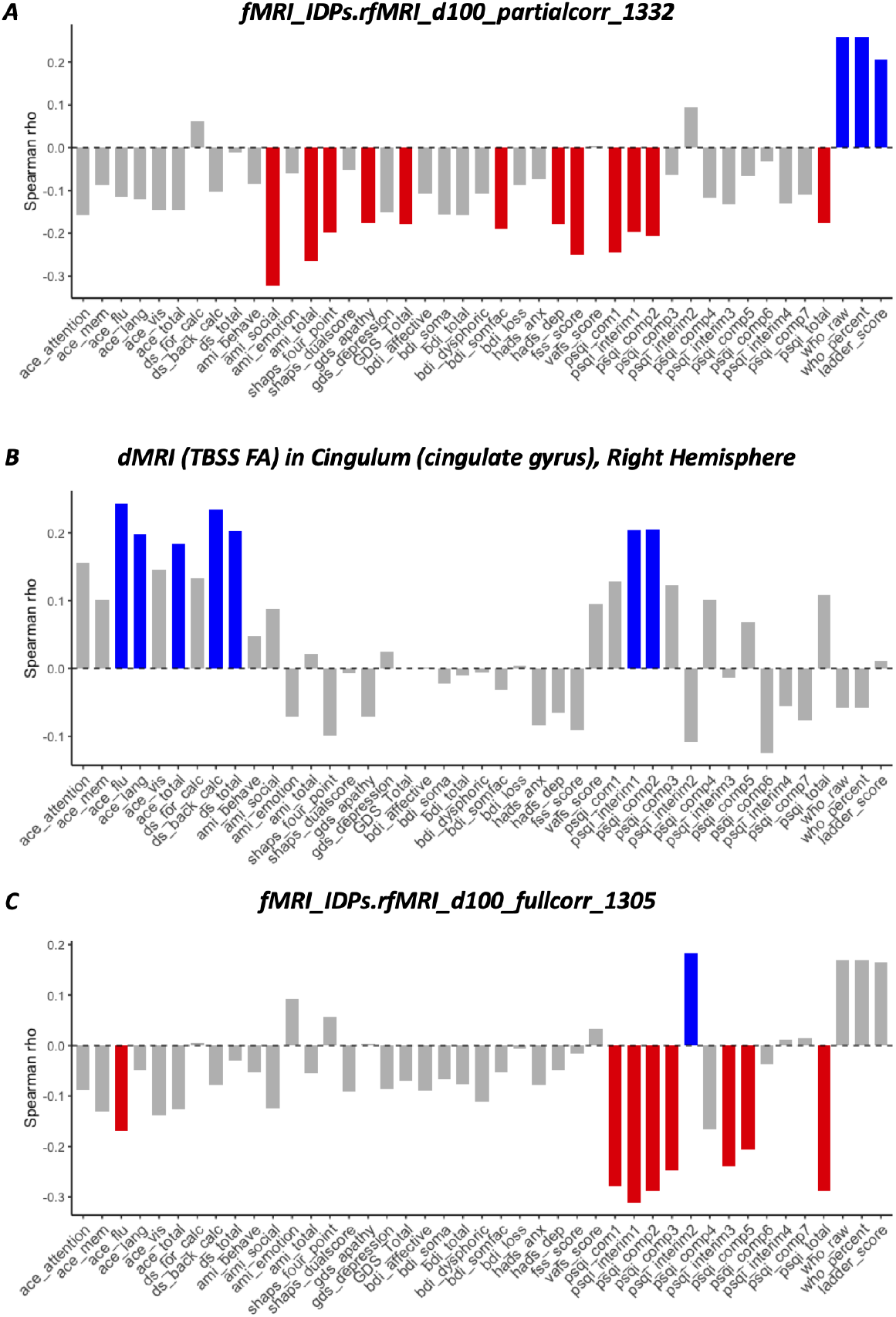
Spearman correlation coefficient for individual symptom measures for key significant IDPs from Fig.5 (panel A) and from Fig.6 (panels B and C). Bars representing symptoms significantly associated with an IDP (uncorrected p*<*0.05) are coloured red or blue according to the direction of the association, whereas bars representing symptoms with non-significant associations are coloured grey.

**Fig. S8.**
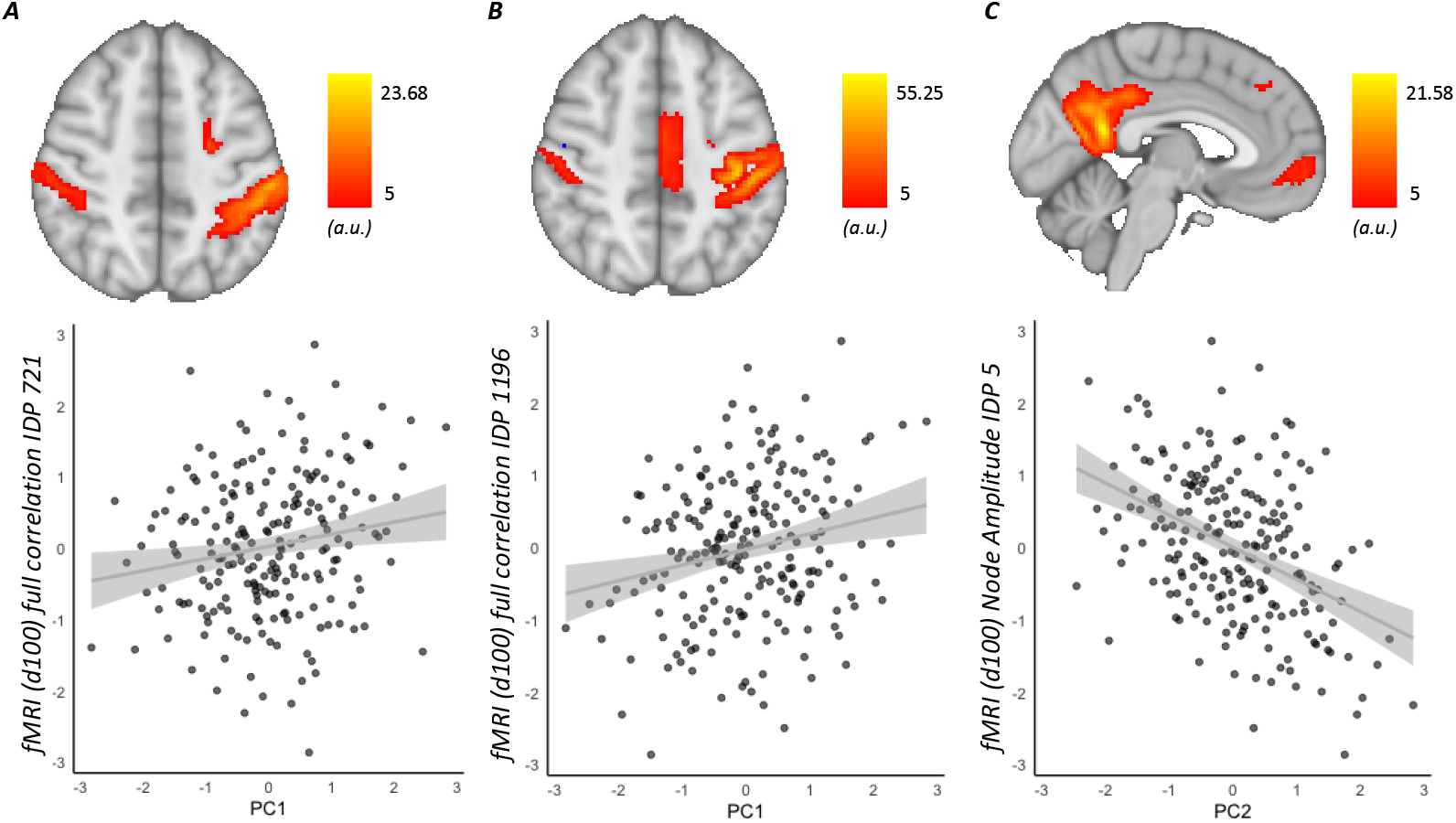
Additional transdiagnostic symptom-associated phenotypes. Associations between resting-state fMRI imaging-derived phenotypes (IDPs) and symptom dimensions derived from principal components (PC1 and PC2). For each association (A–C), the upper panel depicts the spatial distribution of the IDP, and the lower panel shows the corresponding IDP–symptom association. Points represent individual participants.

